# Spatial prediction of COVID-19 pandemic dynamics in the United States

**DOI:** 10.1101/2022.03.27.22271628

**Authors:** Cigdem Ak, Alex D. Chitsazan, Mehmet Gönen, Ruth Etzioni, Aaron J. Grossberg

**Affiliations:** Cancer Early Detection Advanced Research Center, Knight Cancer Institute, Oregon Health & Science University, 2720 S Moody Ave, Portland, OR 97201, USA; Department of Industrial Engineering, College of Engineering, Koç University, Rumelifeneri Yolu, 34450 Sarıyer, İstanbul, Turkey; Program in Biostatistics, Division of Public Health Sciences, Fred Hutchinson Cancer Research Center, 1100 Fairview Ave N, Seattle, WA 98109, USA; Brenden Colson Center for Pancreatic Care, Oregon Health & Science University, 2730 S Moody Ave, Portland, OR 97201, USA; Department of Radiation Medicine, Knight Cancer Institute, Oregon Health & Science University, 3181 SW Sam Jackson Park Road, Portland, OR 97239, USA

## Abstract

**Background:** The impact of COVID-19 across the United States has been heterogeneous, with some areas demonstrating more rapid spread and greater mortality than others. We used geographically-linked data to test the hypothesis that the risk for COVID-19 is spatially defined and sought to define which features are most closely associated with elevated COVID-19 spread and mortality.

**Methods:** Leveraging geographically-restricted social, economic, political, and demographic information from U.S. counties, we developed a computational framework using structured Gaussian processing to predict county-level case and death counts during both the initial and the nationwide phases of the pandemic. After identifying the most predictive spatial features, we applied an unsupervised clustering algorithm, topic modelling, to identify groups of features that are most closely associated with COVID-19 spread.

**Findings:** We found that the inclusion of spatial features modeled case counts very well, with overall Pearson’s correlation coefficient (PCC) and R^2^of 0.96 and 0.84 during the initial phase and 0.95 and 0.87, respectively, during the nationwide phase. The most frequently selected features were associated with urbanicity and 2020 presidential vote margins. When trained using death counts, models revealed similar performance metrics, with the addition of aging metrics to those most frequently selected. Topic modeling showed that counties with similar socioeconomic and demographic features tended to group together, and some feature sets were associated with COVID-19 dynamics. Unsupervised clustering of counties based on these topics revealed groups of counties that experienced markedly different COVID-19 spread.

**Interpretation:** Spatial features explained most of the variability in COVID-19 dynamics between counties. Topic modeling can be used to group collinear features and identify counties with similar features in epidemiologic research.

## Introduction

The COVID-19 pandemic is an unprecedented global health crisis that has infected over 160 million people and taken approximately 4.8 million lives worldwide as of October 4, 2021.^1^ In the United States, spread of COVID rapidly outstripped public health systems, leading to an extremely deadly and widespread pandemic. Even after the initial surge of cases, the nation’s struggles to control disease spread were underscored by ongoing limitations in the availability of personal protective equipment, testing, intensive care unit beds, ventilators, and eventually vaccines. Long incubation period and propensity for asymptomatic spread mean that reactive measures are likely to be too late to quell widespread infection. Targeting intervention to areas at greatest risk for spread in future pandemics could provide a means of suppressing hot spot formation and flattening the pandemic curve.

A range of intersecting biological, demographic, and socioeconomic factors determine susceptibility to COVID-19.^2-4^ These factors vary significantly across areas, and often reflect the structural inequities in the society. Spatial analysis employing Geographical Information Systems (GIS), in which data are layered upon spatial coordinate information, allows researchers to interrogate associations between these factors and COVID-19 pandemic dynamics within and between geographically-defined regions. Research at the county level is well suited to understanding spatial features associated with the pandemic, as COVID-19 spread depends upon proximity, and public health interventions and resources are generally organized at the county level. Studies utilizing GIS reported that, among counties in the United States, measures of income inequality, poverty, urbanity, poor healthcare access, and increased proportion of non-white individuals are associated with COVID-19 incidence and death.^5-8^ Similarly, in England, relative humidity and hospital accessibility are negatively related to COVID-19 mortality rate, whereas percent of Asians, percent of Blacks, and unemployment rate are positively related to COVID-19 mortality rate.^9^

In this study we build upon these known associations with the goal of developing more accurate predictions that capture the heterogeneity in associations between spatial structure and features and compare them across different temporal phases of the pandemic. We curated a large dataset of GIS-tagged demographic, socioeconomic, and political data and utilized structured Gaussian processes (SGP), a machine learning approach to develop dynamic prediction models of localized COVID-19 case and death counts. We applied this approach to both the initial spread of COVID-19 across the US and the dramatic expansion of infections during autumn, 2020, when the virus was ubiquitous, allowing for a direct comparison of factors driving disease dynamics during each epoch of the pandemic. Because many of the most prognostic factors are geographically restricted, we hypothesized that they serve as surrogates for other unmeasured county characteristics. We therefore explored whether counties could be grouped by collinear spatial features to predict those counties with the greatest COVID-19 case burden.

## Results

### A. Analysis of Initial Phase Dynamics

We first sought to define the spatial features that predicted the initial rise in cases, defined here as the 30 days following the first confirmed case in each county. To do so, we trained an SGP regression algorithm on a random sampling of two-thirds of the counties in each state (Figure 1A). For each state, a different set of features were identified to model the dynamics of case spread in each county. We chose to restrict feature selection across counties at the state level, because this represents the main political division at which implementation and timing of mitigation measures and other policies were applied. These state-by-state models were used to generate case predictions in the remaining one-third of “unseen” test counties (Figure 1B), and then compared to the observed case counts in these counties (Figure 1C) to evaluate model performance. Features selected for the prediction models in each state are shown in Figure 1D. The top three most frequently selected features across all states were **Rural-urban continuum code** (higher is more rural), **Vote difference 2020** (%Biden [D] - %Trump [R]), **and urban influence code** (higher is more rural), all of which negatively correlated with case counts (Figure 1E). The next three most frequently selected features—**Total households, total population and domestic migration rate** (net of inmigration - outmigration)—are positively correlated with case counts and reflect the known strong association between population and COVID-19 spread.^10,11^ The remaining top predictive features reflected the importance of health insurance, education, race, income, and population density in predicting case growth. The overall Pearson’s correlation coefficient (PCC) and the proportion of variance explained (R^2^) of this model applied across counties were 0.96 and 0.84, respectively (Figure 1F). Model performance varied across states, with a median PCC of 0.98 [Range 0.74-1] and a median R^2^of 0.94 [Range 0.07-0.99] (Figure 1G). R^2^ was greater than 0.90 in the majority of states, demonstrating that the models built on spatial features could account for most of the variance in case counts.

**Figure 1.**
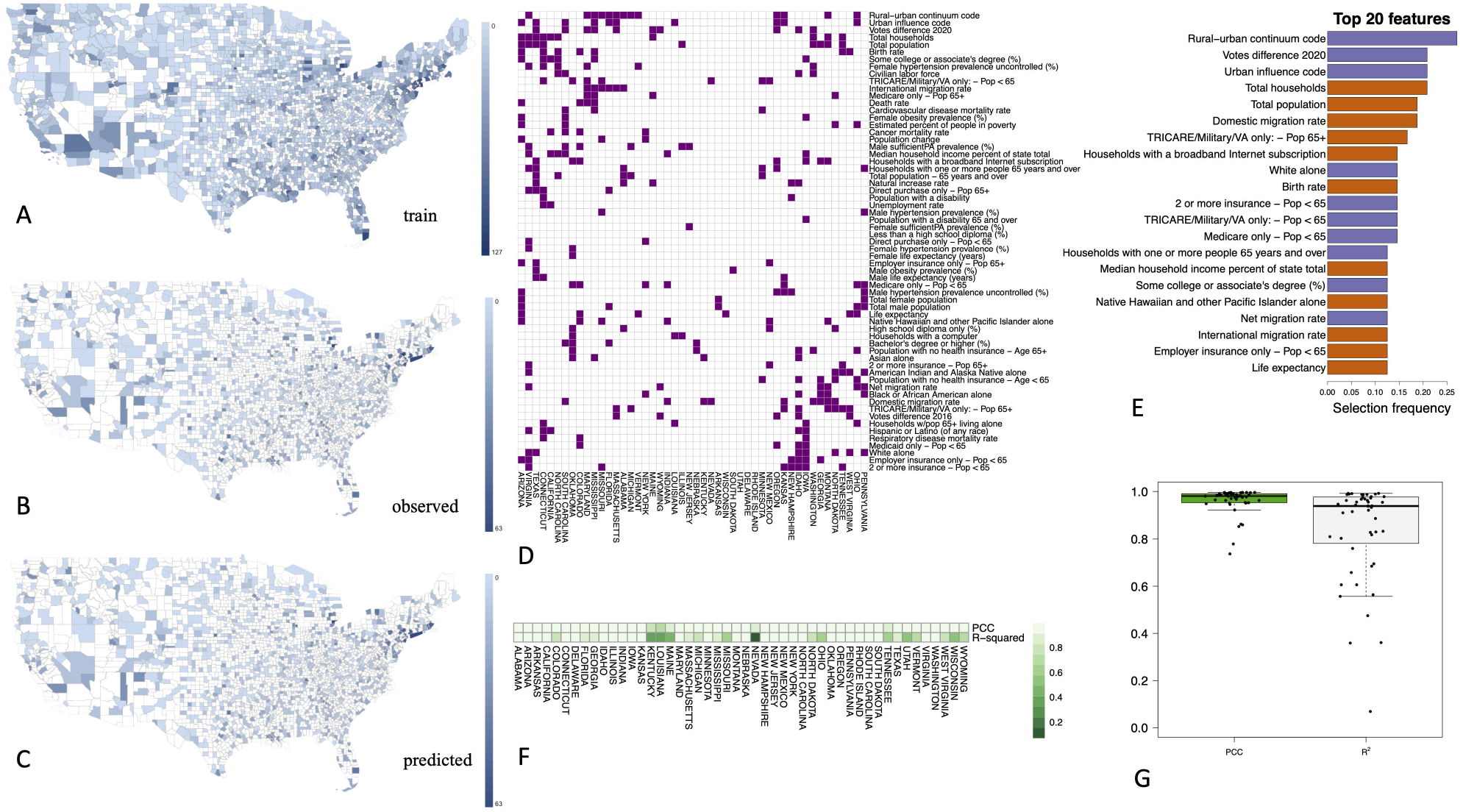
Spatial modeling of case dynamics during initial phase of pandemic. Blue shade indicates observed cases over first 30 days in counties used for model training (A) and testing (B), with predicted case counts in test counties shown in (C). Cases were aggregated over 30 days in each county in the maps. (D) Predictive features selected for modeling in each state. (E) The most predictive top 20 features selected overall by the algorithm for the initial phase. Purple-colored features are negatively correlated with case counts and the orange-colored features are positively correlated with the case counts. (F) PCC and □^2^ values of the predictive models on a state-by-state level. (G) PCC and □^2^ values of all predictions, shown as a box blot.

We then applied an identical approach to generate a spatial model utilizing COVID-19-associated deaths over the first 30 days following the first death in each county as the dependent variable (Supplementary Figure 1). Consistent with prior reports, the features most frequently selected to predict deaths included measures of advanced age and non-white race.^12,13^ **Vote difference 2020** remained the second most frequently selected feature to predict deaths.

### B. Analysis of the Nationwide Phase Dynamics

We next extended our analysis to a later phase of the pandemic, commonly called the “third wave”, which we defined as the period between September 11, 2020, when national case counts were at a local nadir, and March 21, 2021, which marked the next local nadir. In contrast with the initial rise, during this phase the SARS CoV-2 virus was circulating in nearly all counties, testing was more broadly available, and there was a better understanding of modes of spread (droplets and aerosols) and effective mitigation measures, including distancing and masking. Case counts in training counties, predicted case counts in test counties, and observed case counts in test counties are shown in Figure 2A-C, and feature selection for the models derived in each state is shown in Figure 2D. The results largely recapitulated those from the initial phase, with **Urban influence code, Vote difference 2020, Total households, and Total population** the most frequently selected features across all states (Figure 2E). The model again demonstrated a very strong PCC of 0.95 with a R^2^of 0.87, although the model underestimated significant case growth across a subpopulation of counties (Figure 2F). Across states, the model median PCC was 0.98 [Range 0.60-0.99] and the median R^2^was 0.95 [Range 0.20-0.99] (Figure 2G). We generated an independent model to predict deaths during the nationwide phase (Supplementary Figure 2). Because the time interval included both a nationwide rise and fall in cases, which could be governed by different spatial factors, we repeated the models over the rising phase alone, from September 11, 2020 to January 1, 2021. The most frequently selected features during this interval closely reflected those selected over the full epidemic curve, although total female population was selected more frequently in the models predicting deaths over the rising phase (Supplementary Figures 3A-B).

**Figure 2.**
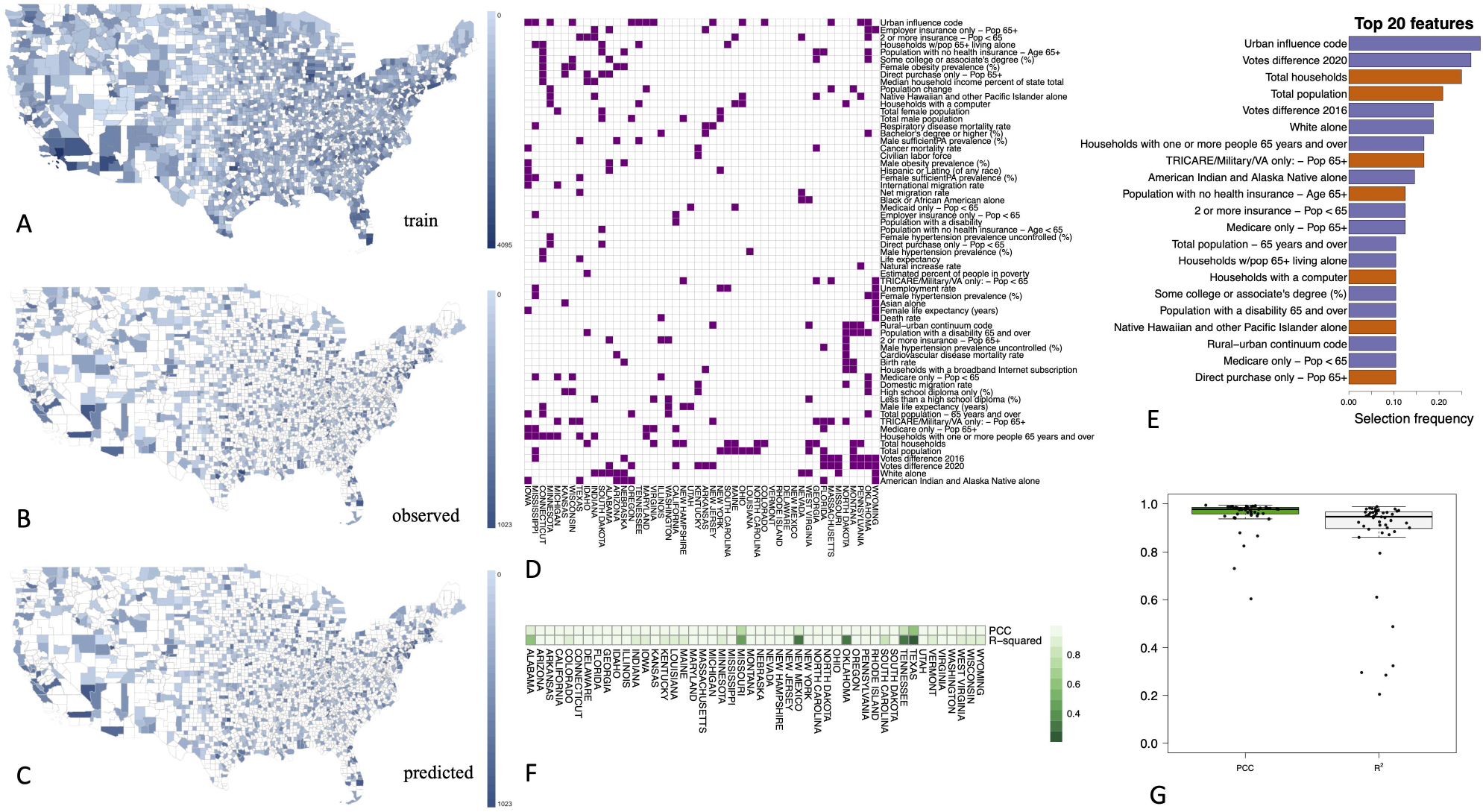
Spatial modeling of case dynamics during nationwide phase of pandemic. Blue shade indicates observed cases over first 30 days in counties used for model training (A) and testing (B), with predicted case counts in test counties shown in (C). Cases were aggregated over the time period after September, 11, 2020 until March 21, 2021 in each county in the maps. (D) Predictive features selected for modeling in each state. (E) The most predictive top 20 features selected overall by the algorithm for the nationwide phase. Purple-colored features are negatively correlated with case counts and the orange-colored features are positively correlated with the case counts. (F) PCC and □^2^ values of the predictive models on a state-by-state level. (G) PCC and □^2^ values of all predictions, shown as a box blot.

We generated daily case and death count predictions for each week *t* across all counties from April 6, 2020 to March 21, 2021 using the spatial features and case counts up through week *t-1* as an internal validation of the selected features sets (Supplementary Figure 4). Consistent with our other analyses, we found that features associated with population and urbanicity, presidential vote margin, and older age were most frequently included in prediction models (Supplementary Figure 5A-G). State-by-state case and death count predictions based on both the spatial and temporal models described above can be reviewed on interactive maps at https://cigdemak.shinyapps.io/sgp_covid-19/.

### C. Topic modeling and unsupervised cluster analysis reveals high risk counties

One limitation of the spatial prediction models described above is that many features are collinear, so the features selected by the SGP modeling are not always the true driver of case growth. Indeed, sets of features cluster along well described socioeconomic, educational, and health axes (Figure 3). Notably, neither **Vote difference 2020** nor **Vote difference 2016** is strongly correlated with any spatial features, suggesting that political leaning of a county is an independent risk factor for COVID-19 spread. Furthermore, the features selected in the models are heterogeneous across states, limiting the ability to define “high risk” locales. For that reason, we set out to group counties by sets of collinear spatial features that together are associated with the risk of COVID-19 spread. To address this, we leveraged a topic modeling (TM) framework using the Latent Dirichlet Allocation (LDA) algorithm to reduce the dimensionality of the data. Using this approach, we utilized TM to find sets of co-occurring features (words) that can then link counties (documents) to topics.

**Figure 3.**
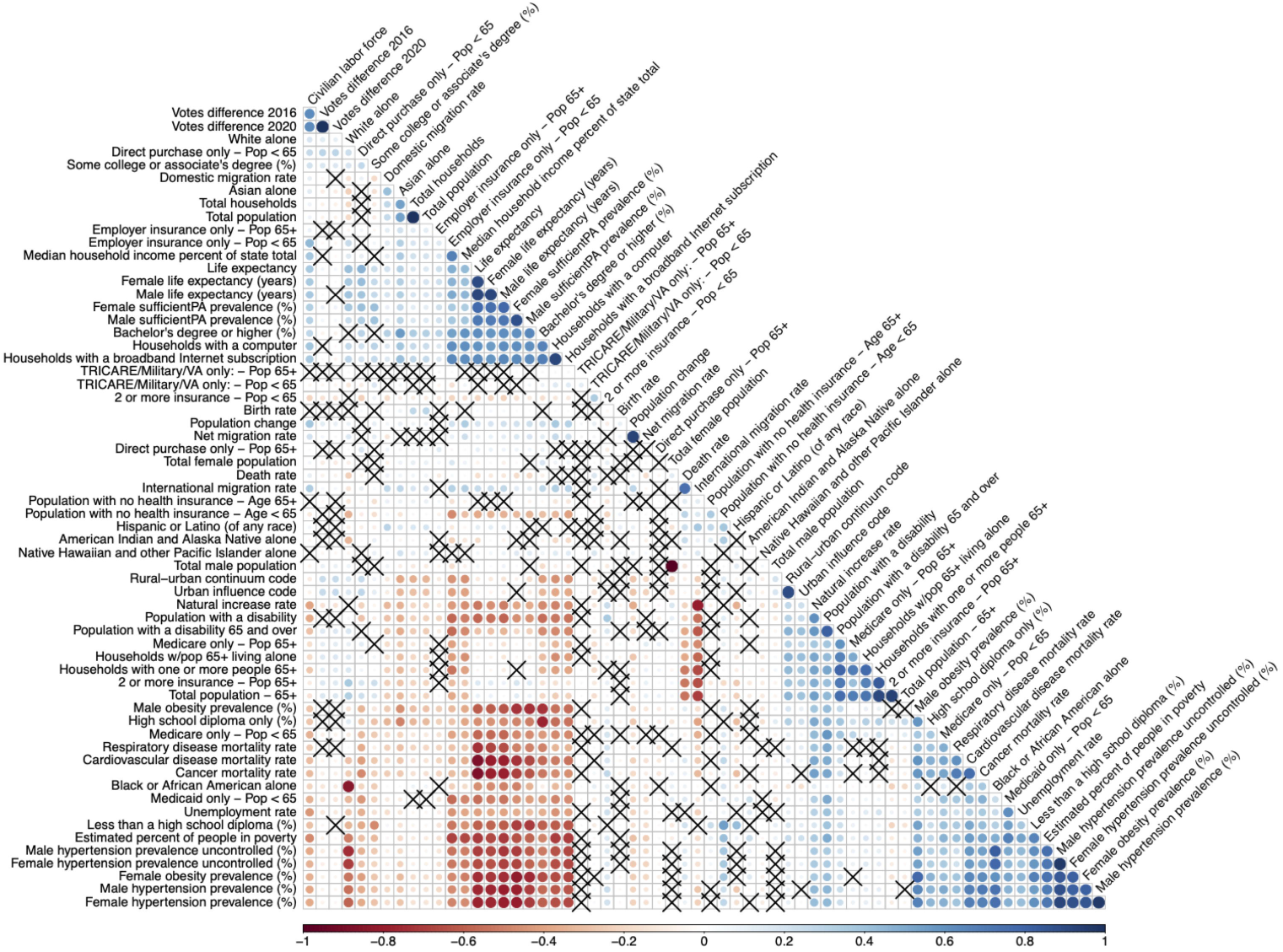
Correlation matrix of the spatial features used in the SGP model. Blue indicates positive correlation and red indicates negative correlation. Shade indicates strength of correlation, per scale shown at bottom of matrix.

The application of TM was able to find sets of collinear features that score each county and feature association to each topic. The top features contributing to each topic are shown in Figure 4A. Topics grouped together many geographically similar counties (Figure 4A), such as topics 2 and 3 which occurred largely in the South and Midwestern regions of the USA respectively. TM also grouped geographically remote but demographically similar counties, such as topic 8, which largely showed features associated with low socioeconomic status. Notably, vote differences were not a primary contributor to any topic, consistent with the low correlation between political orientation and the other features in our dataset. To see how topics related to COVID-19 spread, we looked at the relationship between COVID cases/deaths and topic scores by plotting topic scores against quintiles of cases or deaths for each phase in the. pandemic. Several topics showed correlations with cases and deaths (Figures 4B and 4C). For example, topic 8 (e.g., Less than high school diploma, Percent of people in poverty, Households with supplemental security Income, Medicaid) correlated positively with deaths during the nationwide phase (Figure 4C). Topic 10, which has high feature score contributions from higher education and access to services, showed a negative correlation to the death rate (Figure 4C).

**Figure 4.**
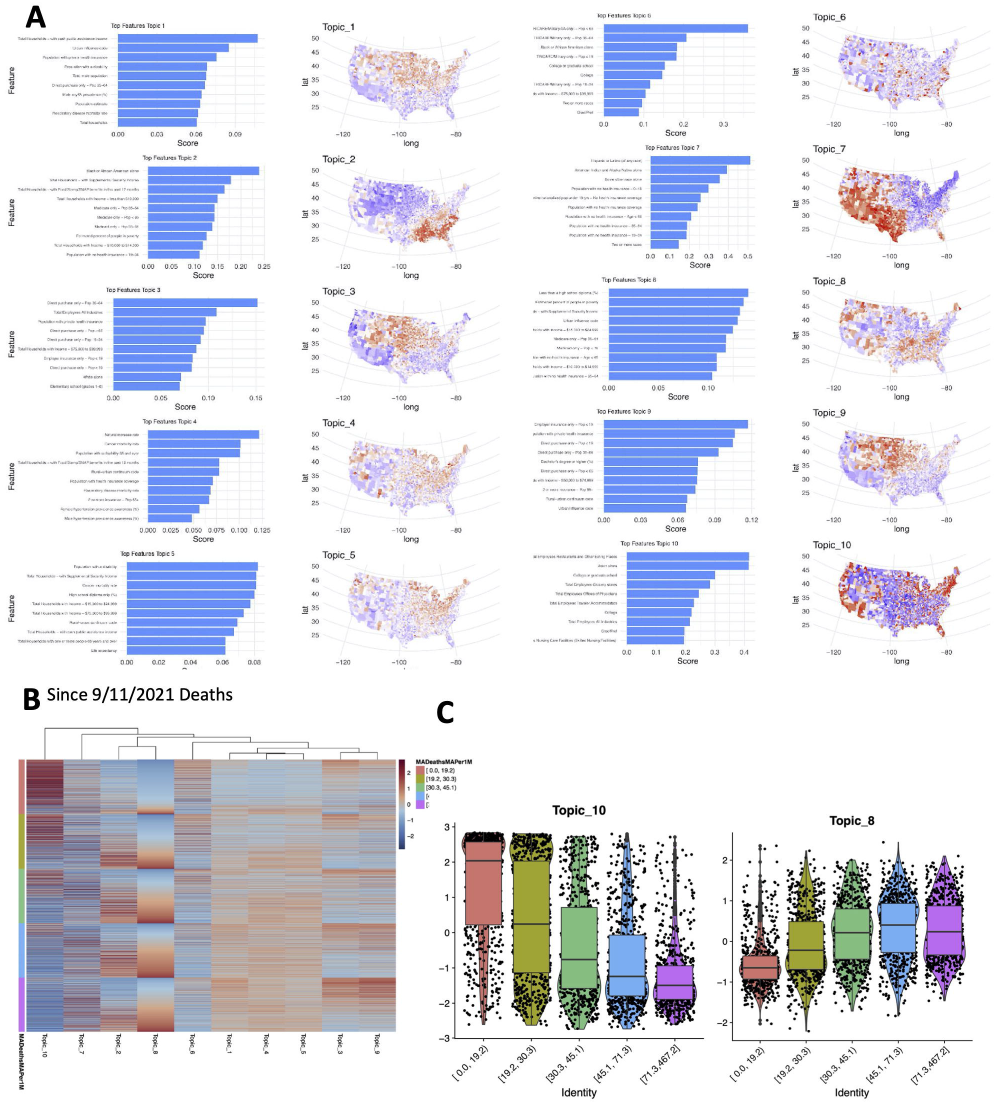
Topic modeling identifies associations between sets of spatial features and COVID-19 dynamics. (A) z-score normalized topic scores for each county in the US as well as the top 10 feature scores for features associated with each topic. (B) Heatmap of each county z-scored topic score against the mean deaths during the nationwide phase, binned into quintiles. To highlight the relationships between topic scores and deaths, the heatmap is sorted by topic 8. (C) Boxplot of topic scores for each county across death quintiles for exemplar topics 10 and 8, showing negative and positive correlations with death counts, respectively.

We therefore clustered the topics using county topic scores in a Louvain clustering algorithm to segregate discrete groups of counties with similar spatial features (topic contributions). After clustering counties with similar socioeconomic and demographic composition tended to group together (Figure 5A). To highlight the feature and topic contributions of each cluster of counties, Figure 5B shows the mean topic score for each topic within each cluster of counties. For example, Cluster 1 is composed of counties with high scores from topics 1, 3 and 9 and low topic 10 scores. This cluster highlights most of the Midwest region where the largest surge in cases and deaths during the autumn, 2020, period of the nationwide phase of the pandemic occurred (Figure 5C). Clustering further delineated cases from deaths and initial phase from nationwide phase dynamics, highlighting plasticity in the composition of spatial features most associated with COVID risk across the course of the pandemic. Cluster 3, which was geographically restricted to the Southeast US, was associated with high COVID-19 case counts during the initial phase, whereas Cluster 0, restricted to Texas, the lower Midwest, and the Rocky Mountain region, was associated with high COVID-19 spread during the nationwide phase (Figure 5C).

**Figure 5.**
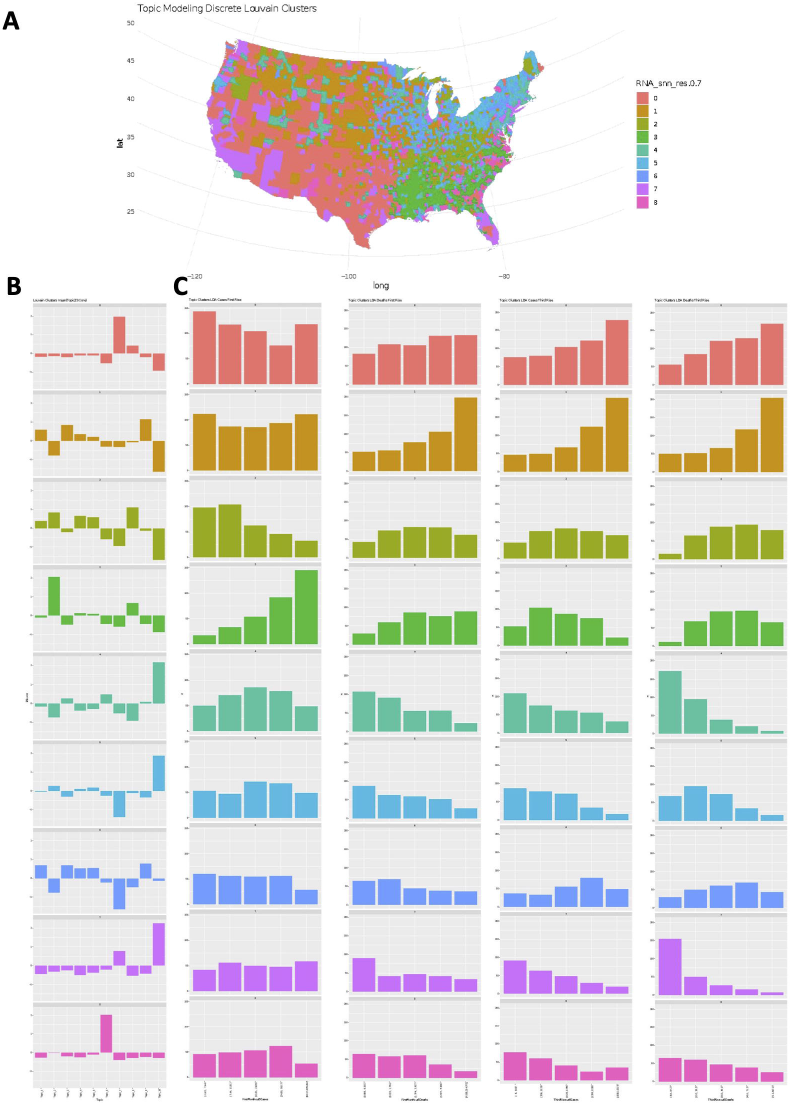
Clustering by topics can identify high and low risk counties. (A) Geographical map of counties and their discrete cluster assignments when topic-county matrix inputted into Louvain clustering. (B) Mean topic score for each topic for each of the 9 clusters of counties. (C) Bar graph of the number of countries within each cluster that fall within each quintile bin of cases and deaths for the initial as well as nationwide phases of the pandemic.

## Discussion

We adopted SGP analysis to generated highly predictive models for case growth, and found that the majority of variance in COVID-19 spread can be explained by the spatial features included in each model. Both case and death counts in each county were most strongly associated with measures of urbanicity, age, and presidential voting margin. We also found that non-white race and measures of socioeconomic status were frequently included in optimal spatial models, recapitulating well-established risk factors for COVID-19 infection and mortality.^14-18^ Extending upon these previously reported models, we show geographic heterogeneity in which factors predict case and mortality across the USA, making it difficult to apply a uniform set of features to identify counties at greatest risk. Because many of the features included in these models are highly correlated, our SGP modeling approach may have obscured stronger effects by diluting selection among collinear features. For example, urban influence code, rural-urban continuum, population density, and total households all describe the urbanicity of a county, yet each individually shows up among the most selected features associated with COVID-19 dynamics, effectively competing for inclusion in the model. Furthermore, these measures of urbanicity were also correlated with the number of individuals over 65 years old, who represent the highest risk cohort for COVID-19 mortality.^19,20^ Correlation analysis also revealed interactions between socioeconomic, health, and racial features, complicating interpretation of the relationships between these features and COVID-19 dynamics.

We therefore sought to identify which combinations of spatial features are most consistently associated with COVID-19 spread across the country using topic modeling to reduce the dimensionality of these data. Although COVID-19 data were not included in the unsupervised groupings, topics were correlated with both cases and deaths. The degree to which a county was represented by a given topic clustered geographically, supporting the utility of this analysis to identify similar places. In accordance with our SGP analysis and prior studies, topics associated with low socioeconomic status correlated with high case and death counts, whereas topics associated with increased wealth and education exhibited an inverse correlation with cases and deaths. By clustering counties according to the overall contribution of topics to their spatial feature set, we were then able to identify those counties across the US that were demographically similar and could show that certain combinations of topics were associated with more case and death burden. These combinations of features likely relate not only to factors that increase rate of spread or mortality, but also adherence and implementation of mitigation measures. Indeed, clusters identified as highest risk largely overlap the National Institute of Environmental Health Sciences’ COVID-19 Pandemic Vulnerability Index,^21^ suggesting that the same spatial features that identified counties with high COVID-19 burden early in the pandemic may drive susceptibility to subsequent waves of infection.

Aside from population metrics, presidential vote margin was the most consistently selected spatial feature in our COVID-19 prediction models. The margin by which a county voted in support of the Republican candidate, Donald Trump, in either the 2016 or 2020 election strongly predicted for more cases and deaths both early and later in the pandemic, adding to prior work revealing this association. Notably, presidential vote margin was not collinear with any other features, suggesting that political orientation represents an independent risk factor for COVID-19 spread. Politics played a prominent role in the US response to the coronavirus pandemic, with mitigation policies and adherence varying widely between areas under Democrat or Republican governance. It is not clear whether this association stems from a “top-down” effect of the administration’s dismissive management and communication approach or reflects growing distrust in science on the ideological right.^22,23^ Indeed, recent work linked partisanship to attitudes about COVID-19 policy and mitigation measures from the beginning of the pandemic, before polarized messaging had developed.^24-26^

The development and implementation of spatially-informed prediction models suffer from several limitations. Our models did not include mitigation measures or vaccine coverage, due in part to inconsistencies in implementation and data availability. The end date for the nationwide phase analysis, March 31, was before vaccine availability was opened to the general public in most states, but differences in vaccine uptake to that point represent a potential confounder. Early case numbers were heavily influenced by low test availability, leading to significant missing data. However, our analyses found similar features predicted case dynamics throughout the pandemic, suggesting that the effect of this missing data may be minimal. Finally, TMand Louvain clustering generate highly overlapping feature sets that may be specific to the breadth of data included. Thus, while spatial analysis provides a powerful predictive tool, the precise effect of each feature or set of features is likely to be context-specific.

In conclusion, we show that spatial features account for the majority of variation in COVID-19 case and death dynamics across the US. Predictive modeling based on combinations of spatial features can identify counties at greatest risk for COVID-19 spread and can be used to direct aggressive mitigation strategies and limited resource pools to these areas. Finally, we show that topic modeling provides a new approach to dimensional reduction in epidemiologic data and may be of value in other datasets with highly collinear variables.

## Methods

We retrieved county-level daily case counts from January 22, 2020 until March 21, 2021 provided by the Center for Systems Science and Engineering at Johns Hopkins University. We extracted county-specific features from the United States Census Bureau and the National Center for Health Statistics population estimates. County-specific features used in this study are shown in Table S1 along with the source information. Boundary shapefile of counties downloaded from TIGER/Line database (https://www.census.gov). We normalized the daily confirmed COVID-19 case and death counts per 100,000 residents and then calculated the 7-day moving average.

### Supervised prediction algorithm: Gaussian process regression

We used Structured Gaussian Processes (SGP) regression algorithm to predict case counts for each county of a given state. SGP allows performing spatiotemporal predictions thanks to the Kronecker multiplication of kernels calculated on spatial and temporal features. After calculating a Gaussian kernel for each spatial and temporal feature, spatial and temporal kernels are combined separately, then combined spatial and temporal kernels are unified with Kronecker multiplication to a larger spatiotemporal kernel, which allows us to make predictions for each given location and time point (Supplementary Figure 4). We calculated a Gaussian kernel for each spatial feature and added it to our feature set only if it improved the prediction quality on the validation set in terms of normalized root mean square error (NRMSE), i.e., forward feature selection. We used the kernel calculated on latitude and longitude of each county by default in the feature selection process.

For the regression algorithm, we designed two different prediction scenarios--spatial prediction and temporal prediction. We performed spatial prediction for (A) initial disease dynamics: the 30 days following the first case in each county and (B) nationwide disease dynamics: the time period between September 11, 2020, when the nationwide rise in cases began and March 21 2021, when the epidemic curve is completed (see Supplementary Figure 5). For both phases, we randomly selected two-thirds of the counties in each state to train our model and predicted case and death counts of the remaining one-third of the counties in each state and repeated the training--testing selection 100 times to eliminate random sampling. In temporal prediction, we are interested in finding case counts in observed locations for a future unseen time period. We predicted daily COVID-19 case counts/death counts for each location at the beginning of each week for a week starting from April 6, 2020, the peak of the first rise, until March 21, 2021. We used the last week of training dataset as the validation dataset to select the spatial features with forward selection method and also to optimize the model’s response noise parameter. Accuracy was assessed using PCC, which shows how well the dynamics of the event counts are captured by the algorithms, and R^2^, which shows the proportion of total variation in outcomes explained by the model. SGP implementation in R is publicly available.^27^

### Unsupervised prediction algorithm: Topic Modeling

TM using the LDA algorithm, in the traditional framework, associates words and documents to topics by linking together co-occurring words in *k*-number of topics, which then can be related to the documents by comparing the relative occurrence of said words in each topic, outputting a topic-word and topic-document distribution. Using this approach, TM finds sets of co-occurring features (words) that can then link counties (documents) to topics. We ran LDA with the package ‘lda’ in R. Total number of topics was found using the rate of perplexity change elbow plot reported by Zhao and colleagues.^28^ To visualize how cases and deaths related to topics, deaths and cases from the initial phase and nationwide phase, as described above, were binned into 5 categorical quintiles of mean cases/100k and deaths/100k and regressed against average topic scores.

### Clustering counties

To group counties together by the relative contributions of topics to each county, we imputed the dimensionally reduced LDA topics into a Louvain clustering algorithm using a resolution of 0.7, which resulted in 9 clusters. Topic contributions were then shown by plotting the average z-score normalized topic scores across all counties within a given cluster. Then to see which clusters had a high incidence of deaths/cases per capita, we plotted a histogram of the number of counties across each quintile.

### Role of the funding source

Sponsors had no role in in study design; in the collection, analysis, and interpretation of data; in the writing of the report; or in the decision to submit the paper for publication.

## Data Availability

All data produced are publicly available online at the sources identified in Table S1.

## Declaration of Interests

We declare no competing interests.

## Acknowledgements

This work was supported by funding from the Cancer Early Detection Advanced Research Center at the Knight Cancer Institute, Oregon Health & Science University (CA, ADC, RE, and AJG) and the National Cancer Institute (K08 CA245188) awarded to AJG.

## Notes

### Competing Interest Statement

The authors have declared no competing interest.

### Author Declarations

All data used in this manuscript are publicly available. We retrieved county-level daily case counts from the Center for Systems Science and Engineering at Johns Hopkins University. We extracted county-specific features from the United States Census Bureau and the National Center for Health Statistics population estimates. County-specific features used in this study are shown in Table S1 along with the source information.

