## Supplementary Materials for "Spatial prediction of COVID-19 pandemic dynamics in the United States"

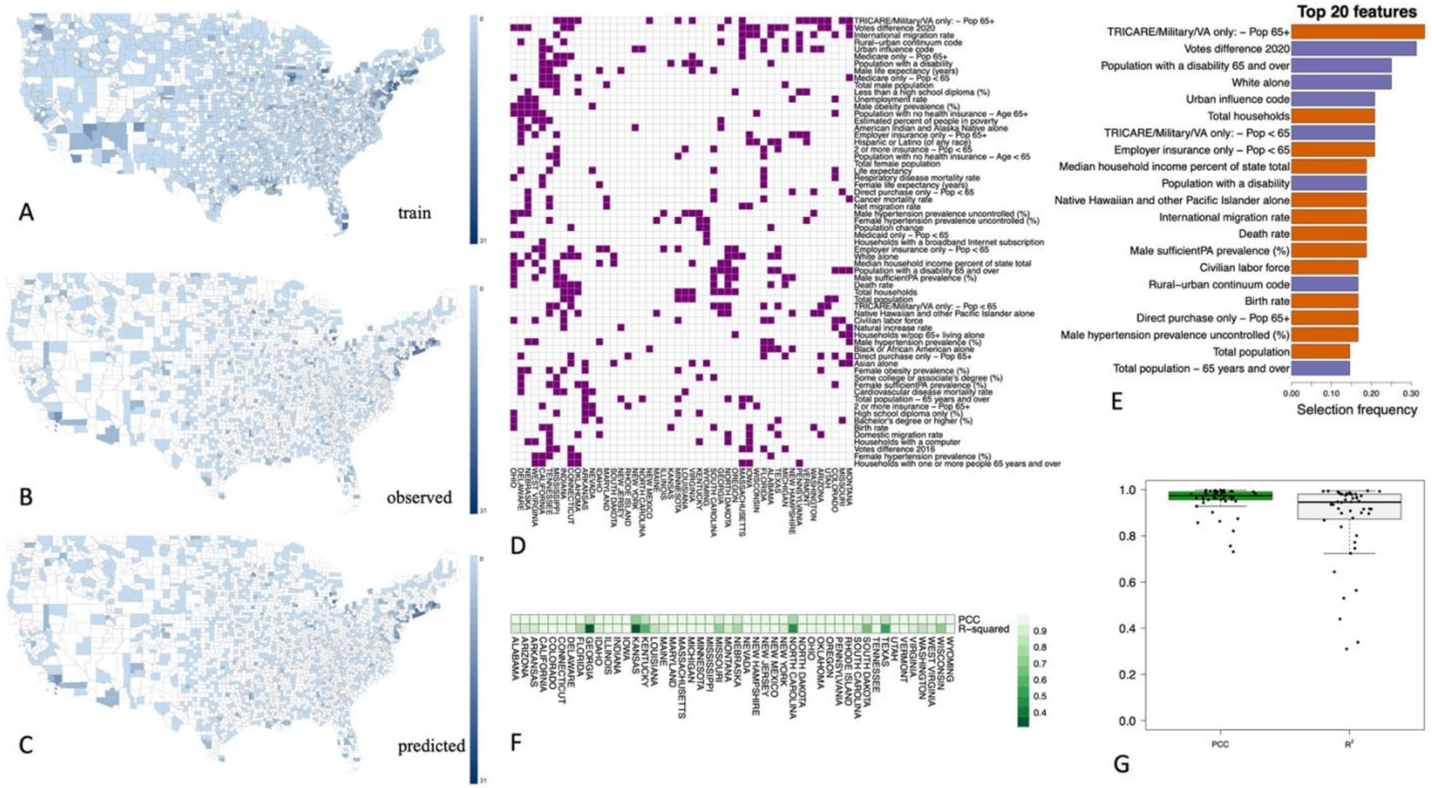


Supplementary Figure 1

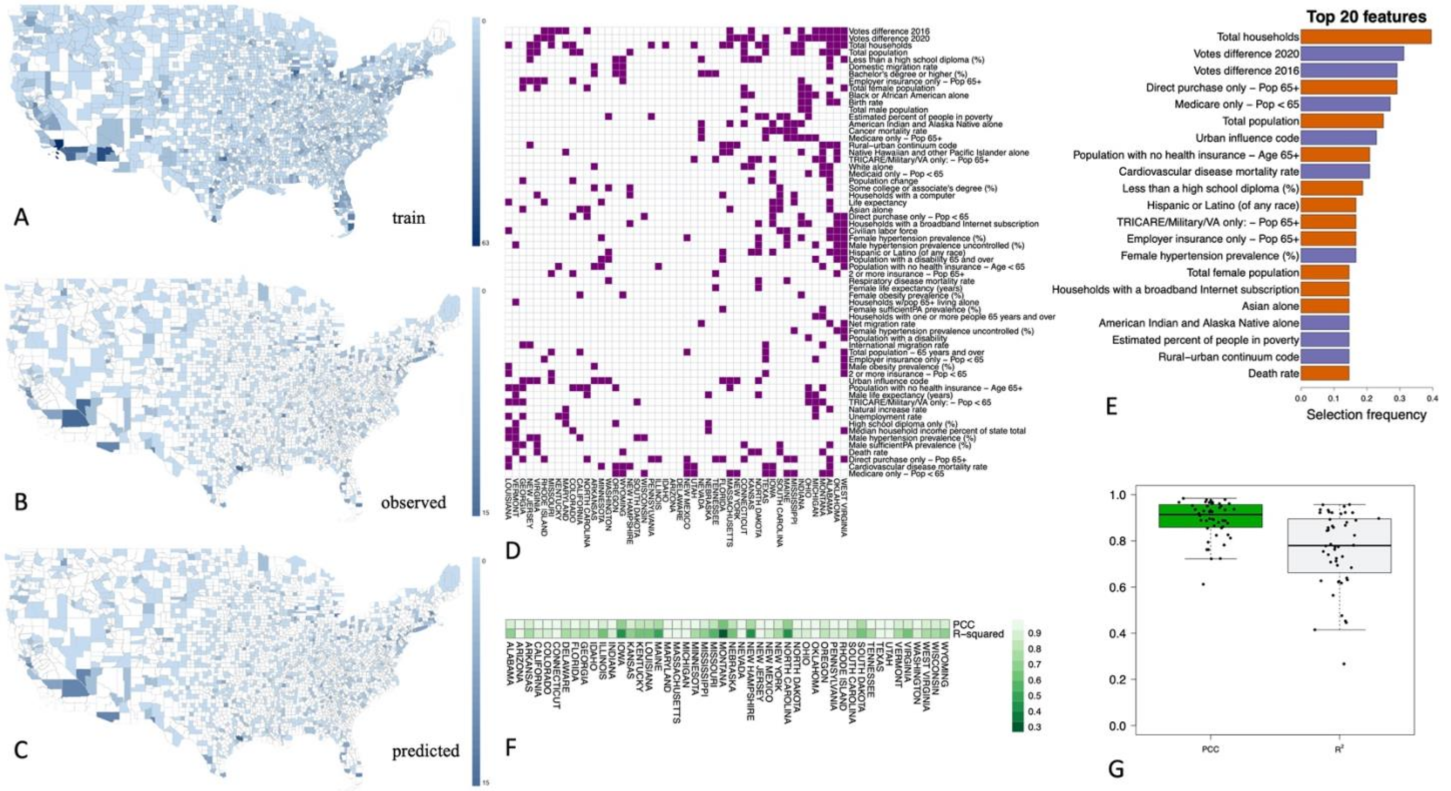


Supplementary Figure 2

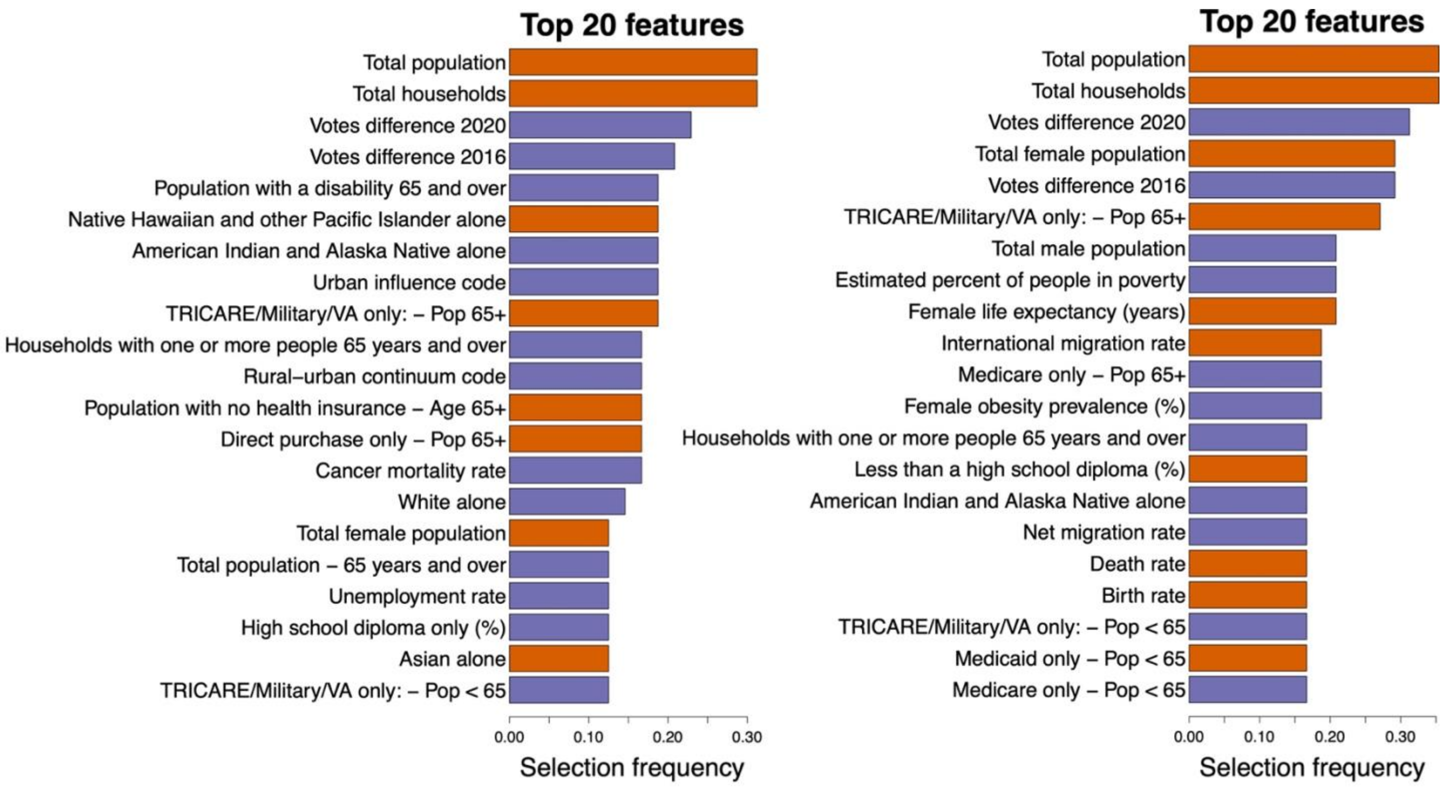


Supplementary Figure 3

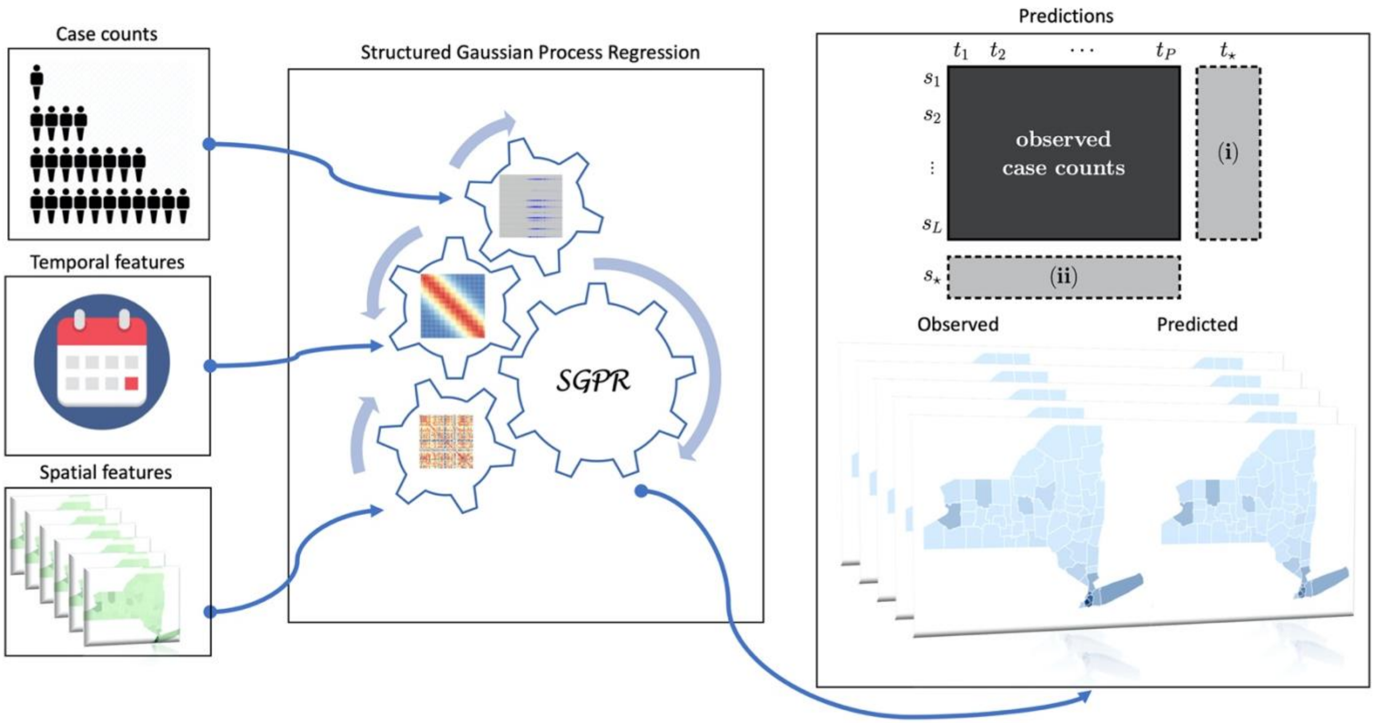


Supplementary Figure 4

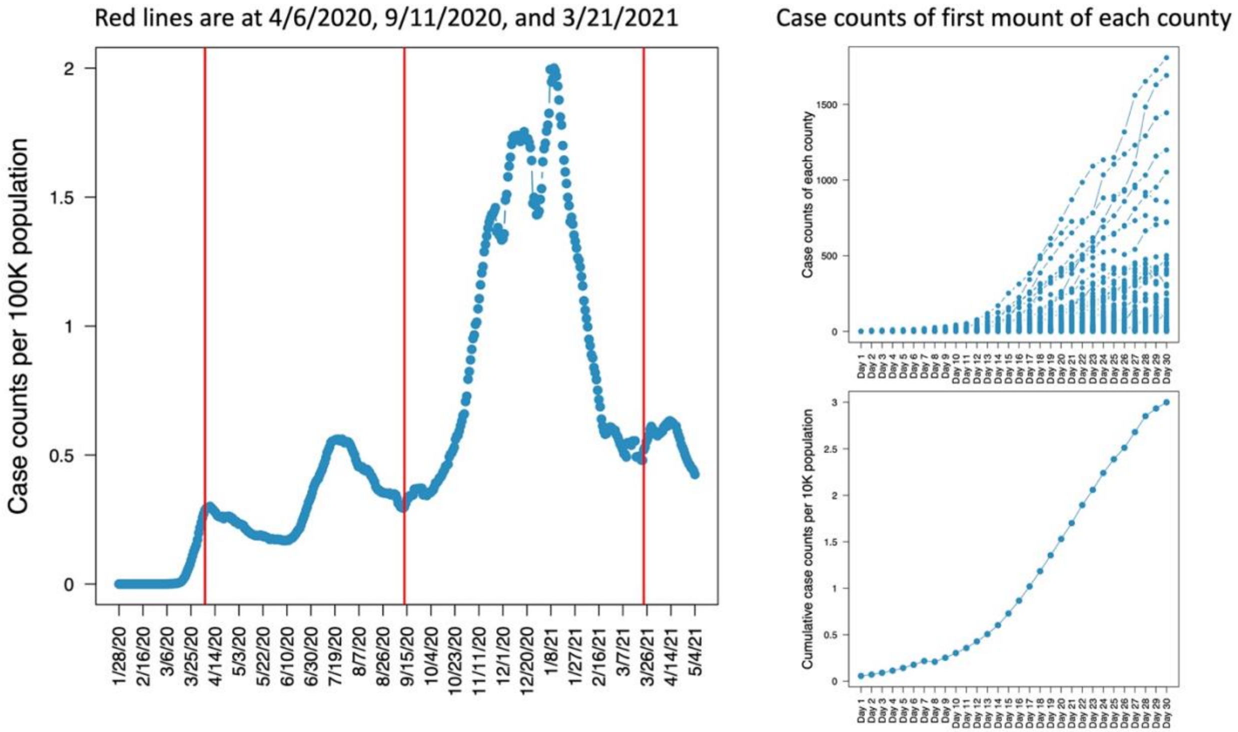


Supplementary Figure 5

**Supplementary Figure Legends**

**Supplementary Figure 1. Spatial modeling of death dynamics during initial phase of pandemic.** Blue shade indicates observed deaths over first 30 days in counties used for model training (A) and testing (B), with predicted death counts in test counties shown in (C). Death counts were aggregated over 30 days in each county in the maps. (D) Predictive features selected for modeling in each state. (E) The most predictive top 20 features selected overall by the algorithm for the initial phase. Purple-colored features are negatively correlated with death counts, and the orange-colored features are positively correlated with death counts. (F) PCC and $R^2$ values of the predictive models on a state-by-state level. (G) PCC and $R^2$ values of all predictions, shown as a box blot.

**Supplementary Figure 2. Spatial modeling of death dynamics during nationwide phase of pandemic.** Blue shade indicates observed deaths over first 30 days in counties used for model training (A) and testing (B), with predicted death counts in test counties shown in (C). Death counts were aggregated over the time period after September, 11, 2020 until March 21, 2021 in each county in the maps. (D) Predictive features selected for modeling in each state. (E) The most predictive top 20 features selected overall by the algorithm for the initial phase. Purple-colored features are negatively correlated with death counts, and the orange-colored features are positively correlated with death counts. (F) PCC and $R^2$ values of the predictive models on a state-by-state level. (G) PCC and $R^2$ values of all predictions, shown as a box blot.

**Supplementary Figure 3.** The top 20 most frequently selected features to predict cases (left) and deaths (right) from September 11, 2020 to January 1, 2021. Purple-colored features are negatively correlated with the death counts, and orange-colored features are positively correlated with the case counts.

**Supplementary Figure 4.** Overview of our predictive computational framework SGP.

**Supplementary Figure 5.** U.S. wide total 7-day moving average case counts per 100,000 population (left). Red lines indicate the dates we selected for analysis of initial and nationwide pandemic dynamics. Case counts in each county (right, top) and cumulative case counts per 10,000 population (right, bottom) during first 30 days following first confirmed case in each county.

**Supplementary Table 1**

| Feature | Source | Description |
|---|---|---|
| Latitude & longitude | USA shape files | Geograpical coordinates of counties |
| Female obesity prevalence | http://ghdx.healthdata.org/us-data | Female obesity prevalence 2011 (%) |
| Male obesity prevalence | http://ghdx.healthdata.org/us-data | Male obesity prevalence 2011 (%) |
| Female anyPA prevalence | http://ghdx.healthdata.org/us-data | Prevalence 2011 (%) |
| Male anyPA prevalence | http://ghdx.healthdata.org/us-data | Prevalence 2011 (%) |
| Female sufficientPA prevalence | http://ghdx.healthdata.org/us-data | Female sufficientPA Prevalence 2011 (%) |
| Male sufficientPA prevalence | http://ghdx.healthdata.org/us-data | Male sufficientPA Prevalence 2011 (%) |
| Cardiovascular disease mortality | http://ghdx.healthdata.org/us-data | Mortality rate 2014 |
| Cancer mortality | http://ghdx.healthdata.org/us-data | Mortality rate 2014 |
| Respiratory disease mortality | http://ghdx.healthdata.org/us-data | Mortality rate 2014 |
| Life expectancy | http://ghdx.healthdata.org/us-data | 2014 |
| Male hypertension prevalence (%) | http://ghdx.healthdata.org/us-data | total hypertension prevalence (among all respondents, percentage of those who reported systolic BP of at least 140mm HG and/or self-reported taking medication) |
| Female hypertension prevalence (%) | http://ghdx.healthdata.org/us-data | total hypertension prevalence (among all respondents, percentage of those who reported systolic BP of at least 140mm HG and/or self-reported taking medication) |
| Male hypertension prevalence self-reported (%) | http://ghdx.healthdata.org/us-data | percentage of respondents who reported being told by a doctor or other healthcare professional that they had hypertension |
| Female hypertension prevalence self-reported (%) | http://ghdx.healthdata.org/us-data | percentage of respondents who reported being told by a doctor or other healthcare professional that they had hypertension |
| Male hypertension prevalence treatment (%) | http://ghdx.healthdata.org/us-data | among prevalent cases, those who report taking prescribed medication |
| Female hypertension prevalence treatment (%) | http://ghdx.healthdata.org/us-data | among prevalent cases, those who report taking prescribed medication |
| Male hypertension prevalence control (%) | http://ghdx.healthdata.org/us-data | among prevalent cases, the percentage of respondents who reported taking prescribed medication and SBP <140 mm Hg |

| | | |
|---|---|---|
| Female hypertension prevalence control (%) | http://ghdx.healthdata.org/us-data | among prevalent cases, the percentage of respondents who reported taking prescribed medication and SBP <140 mm Hg |
| Male hypertension prevalence uncontrolled (%) | http://ghdx.healthdata.org/us-data | percentage of respondents who reported being told by a doctor or other healthcare professional that they had hypertension and SBP of 140 mm Hg or higher |
| Female hypertension prevalence uncontrolled (%) | http://ghdx.healthdata.org/us-data | percentage of respondents who reported being told by a doctor or other healthcare professional that they had hypertension and SBP of 140 mm Hg or higher |
| Male hypertension prevalence awareness (%) | http://ghdx.healthdata.org/us-data | among prevalent cases, the percentage of people who have hypertension and know it |
| Female hypertension prevalence awareness (%) | http://ghdx.healthdata.org/us-data | among prevalent cases, the percentage of people who have hypertension and know it |
| Direct purchase only - Pop 19-34 | https://covid19.census.gov/datasets/ | Number of Population |
| Direct purchase only - Pop 35-64 | https://covid19.census.gov/datasets/ | Number of Population |
| Direct purchase only - Pop 65+ | https://covid19.census.gov/datasets/ | Number of Population |
| Medicare only - Pop < 19 | https://covid19.census.gov/datasets/ | Number of Population |
| Medicare only - Pop 19-34 | https://covid19.census.gov/datasets/ | Number of Population |
| Medicare only - Pop 35-64 | https://covid19.census.gov/datasets/ | Number of Population |
| Medicare only - Pop 65+ | https://covid19.census.gov/datasets/ | Number of Population |
| Medicaid only - Pop < 19 | https://covid19.census.gov/datasets/ | Number of Population |
| Medicaid only - Pop 19-34 | https://covid19.census.gov/datasets/ | Number of Population |
| Medicaid only - Pop 35-64 | https://covid19.census.gov/datasets/ | Number of Population |
| TRICARE/Military only: - Pop < 19 | https://covid19.census.gov/datasets/ | Number of Population |
| TRICARE/Military only: - Pop 19-34 | https://covid19.census.gov/datasets/ | Number of Population |
| TRICARE/Military only: - Pop 35-64 | https://covid19.census.gov/datasets/ | Number of Population |
| TRICARE/Military only: - Pop 65+ | https://covid19.census.gov/datasets/ | Number of Population |
| VA health care only - Pop < 19 | https://covid19.census.gov/datasets/ | Number of Population |
| VA health care only - Pop 19-34 | https://covid19.census.gov/datasets/ | Number of Population |
| VA health care only - Pop 35-64 | https://covid19.census.gov/datasets/ | Number of Population |
| VA health care only - Pop 65+ | https://covid19.census.gov/datasets/ | Number of Population |
| 2 or more insurance - Pop < 19 | https://covid19.census.gov/datasets/ | Number of Population |
| 2 or more insurance - Pop 19-34 | https://covid19.census.gov/datasets/ | Number of Population |

| | | |
|---|---|---|
| 2 or more insurance - Pop 35-64 | https://covid19.census.gov/datasets/ | Number of Population |
| 2 or more insurance - Pop 65+ | https://covid19.census.gov/datasets/ | Number of Population |
| Population with no health insurance - 0-18 | https://covid19.census.gov/datasets/ | Number of Population |
| Population with no health insurance - 19-34 | https://covid19.census.gov/datasets/ | Number of Population |
| Population with no health insurance - 35-64 | https://covid19.census.gov/datasets/ | Number of Population |
| Population with no health insurance - Age 65+ | https://covid19.census.gov/datasets/ | Number of Population |
| Civilian noninstitutionalized pop - With health coverage | https://covid19.census.gov/datasets/ | Number of Population |
| Total civilian noninstitutionalized pop - With health insurance coverage | https://covid19.census.gov/datasets/ | Number of Population |
| Total civilian noninstitutionalized pop - With private health insurance | https://covid19.census.gov/datasets/ | Number of Population |
| Total civilian noninstitutionalized pop - With public coverage | https://covid19.census.gov/datasets/ | Number of Population |
| Total civilian noninstitutionalized pop - No health insurance coverage | https://covid19.census.gov/datasets/ | Number of Population |
| Civilian noninstitutionalized pop under 19 yrs | https://covid19.census.gov/datasets/ | Number of Population |
| Total civilian noninstitutionalized pop under 19 yrs - No health insurance coverage | https://covid19.census.gov/datasets/ | Number of Population |
| Total civilian noninstitutionalized pop - with a disability | https://covid19.census.gov/datasets/ | Number of Population |
| Total civilian noninstitutionalized pop - with a disability 65 and over | https://covid19.census.gov/datasets/ | Number of Population |
| Total households with a computer | https://covid19.census.gov/datasets/ | Number of total households |
| Total households with a broadband Internet subscription | https://covid19.census.gov/datasets/ | Number of total households |
| Less than a high school diploma | https://www.ers.usda.gov/data-products/county-level-data-sets/download-data/ | Percent of adults, 2014-18 |
| High school diploma only | https://www.ers.usda.gov/data-products/county-level-data-sets/download-data/ | Percent of adults, 2014-19 |
| Some college or associate's degree | https://www.ers.usda.gov/data-products/county-level-data-sets/download-data/ | Percent of adults, 2014-20 |

| | | |
|---|---|---|
| Bachelor's degree or higher | https://www.ers.usda.gov/data-products/county-level-data-sets/download-data/ | Percent of adults, 2014-21 |
| Population estimate | https://www.ers.usda.gov/data-products/county-level-data-sets/download-data/ | Population estimate 2019 |
| Population change | https://www.ers.usda.gov/data-products/county-level-data-sets/download-data/ | Number of population change 2019 |
| Births | https://www.ers.usda.gov/data-products/county-level-data-sets/download-data/ | Birth rate 2019 |
| Deaths | https://www.ers.usda.gov/data-products/county-level-data-sets/download-data/ | Death rate 2019 |
| Natural increase | https://www.ers.usda.gov/data-products/county-level-data-sets/download-data/ | Natural increase rate 2019 |
| International migration | https://www.ers.usda.gov/data-products/county-level-data-sets/download-data/ | International migration rate 2019 |
| Domestic migration | https://www.ers.usda.gov/data-products/county-level-data-sets/download-data/ | Domestic migration rate 2019 |
| Net migration | https://www.ers.usda.gov/data-products/county-level-data-sets/download-data/ | Net migration rate 2019 |
| Residual | https://www.ers.usda.gov/data-products/county-level-data-sets/download-data/ | Residual 2019 |
| Rural-urban continuum code | https://www.ers.usda.gov/data-products/county-level-data-sets/download-data/ | The 2013 Rural-Urban Continuum Codes form a classification scheme that distinguishes metropolitan counties by the population size of their metro area, and nonmetropolitan counties by degree of urbanization and adjacency to a metro area. |

| | | |
|---|---|---|
| Urban influence code | https://www.ers.usda.gov/data-products/county-level-data-sets/download-data/ | The 2013 Urban Influence Codes form a classification scheme that distinguishes metropolitan counties by population size of their metro area, and nonmetropolitan counties by size of the largest city or town and proximity to metro and micropolitan areas. |
| White alone | https://covid19.census.gov/datasets/ | Number of Total Population |
| Black or African American alone | https://covid19.census.gov/datasets/ | Number of Total Population |
| American Indian and Alaska Native alone | https://covid19.census.gov/datasets/ | Number of Total Population |
| Asian alone | https://covid19.census.gov/datasets/ | Number of Total Population |
| Native Hawaiian and other Pacific Islander alone | https://covid19.census.gov/datasets/ | Number of Total Population |
| Some other race alone | https://covid19.census.gov/datasets/ | Number of Total Population |
| Two or more races | https://covid19.census.gov/datasets/ | Number of Total Population |
| Not Hispanic or Latino | https://covid19.census.gov/datasets/ | Number of Total Population |
| Hispanic or Latino (of any race) | https://covid19.census.gov/datasets/ | Number of Total Population |
| Nursery, preschool | https://covid19.census.gov/datasets/ | Number of Population Enrollment in |
| Kindergarten | https://covid19.census.gov/datasets/ | Number of Population Enrollment in |
| Grade 1-4 | https://covid19.census.gov/datasets/ | Number of Population Enrollment in |
| Grade 5-8 | https://covid19.census.gov/datasets/ | Number of Population Enrollment in |
| Grade 9-12 | https://covid19.census.gov/datasets/ | Number of Population Enrollment in |
| College | https://covid19.census.gov/datasets/ | Number of Population Enrollment in |
| Grad/Prof | https://covid19.census.gov/datasets/ | Number of Population Enrollment in |
| Elementary school (grades 1-8) | https://covid19.census.gov/datasets/ | Number of Population Enrollment in |
| High school (grades 9-12) | https://covid19.census.gov/datasets/ | Number of Population Enrollment in |
| College or graduate school | https://covid19.census.gov/datasets/ | Number of Population Enrollment in |
| Total households | https://covid19.census.gov/datasets/ | Number of Total Households |
| Civilian labor force | https://www.ers.usda.gov/data-products/county-level-data-sets/download-data/ | Number, 2019 |
| Employed | https://www.ers.usda.gov/data-products/county-level-data-sets/download-data/ | Rate 2019 |
| Unemployed | https://www.ers.usda.gov/data-products/county-level-data-sets/download-data/ | Rate 2019 |

| | | |
|---|---|---|
| Median household income | https://www.ers.usda.gov/data-products/county-level-data-sets/download-data/ | Number, 2019 |
| Median household income percent of state total | https://www.ers.usda.gov/data-products/county-level-data-sets/download-data/ | Number, 2020 |
| Votes difference | https://github.com/tonmcg/US_County_Level_Election_Results_08-20 | Absolute difference between Republican votes - Democrat votes |
| Estimated percent of people in poverty | https://www.ers.usda.gov/data-products/county-level-data-sets/download-data/ | Estimated percent of people of all ages in poverty 2018 |

**Supplementary Table Legends**

**Supplementary Table 1.** Full list of features and sources used in the development of spatial models of COVID-19 dynamics.
